# Sentinel Surveillance for Pediatric Bacterial Meningitis in a Tertiary-Level Pediatric Hospital in Colombia, 2016–2023

**DOI:** 10.64898/2026.03.25.26348800

**Authors:** Adriana Bautista, Germán Camacho-Moreno, Daniela Jerez, María del Pilar Perdomo Rojas, Jaime Moreno, Luz Yanet Maldonado, Yenny Marcela Elizalde Rodriguez, Olga Sanabria, Jacqueline Palacios, Jaid Constanza Rojas Sotelo, María Cristina Duarte, Eliana Sabogal, Karen Jimenez, Carolina Duarte

## Abstract

**Introduction:** Bacterial meningitis (BM) is a common bacterial infection of the central nervous system, and its incidence in children varies by age, with the highest rates observed in infants younger than two months old.

**Objective:** To describe the etiology, epidemiology, and clinical presentation of children under 5 years of age with BM at HOMI between 2016 to 2023.

**Materials and methods:** Descriptive study of children under 5 years of age with suspected BM. Probable cases were those with CSF results consistent with BM. Confirmed cases had a positive CSF culture or blood culture for a bacterial pathogen or a positive molecular test for a bacterium in the CSF. Demographic variables, incidence of BM per year, mortality, and sequelae among survivors were analyzed.

**Results:** A total of 527 suspected cases of BM were evaluated. Of these, 22.8% (120/527) were classified as probable cases and 13.1% (69/527) as confirmed cases. Children under 2 years of age accounted for 37.2% of probable cases and 78.2% of confirmed cases. Among confirmed cases, the most frequent symptoms were fever (98.3%), altered consciousness (39.1%), seizures (36.2%), and lethargy (27.5%). The mortality rate was 11.6% (8/69), and the mean hospital stay among patients with BM was 24.5 days. *Streptococcus pneumoniae* was identified in 26.1% of confirmed cases, with most isolates belonging to serotypes not included in PCV10. *Haemophilus influenzae* accounted for 17.4% of cases, of which 77.7% were serotype b. *Neisseria meningitidis* represented 10.1% of cases, and 60% of these were serogroup C. Other pathogens were identified in 49.1% of patients. **Conclusion**: Sentinel surveillance makes it possible to measure the impact of public health interventions and evaluate the impact of vaccines already used.

## INTRODUCTION

Over the past three decades, substantial progress has been made in the prevention and treatment of community acquired bacterial meningitis; however, the global burden of the disease remains high. The incidence rates range from approximately 0.9 to 80 per 100,000 individuals per year in high-income and low-income countries, respectively (1). In low-income countries, BM has a mortality rate of up to 54% and up to 24% of survivors develop chronic neurological sequelae such as hearing loss or focal neurological deficits (1).

The distribution of pathogens causing BM varies according to age, immune status, geographical region, and the impact of conjugate vaccines against the most common bacteria (2). In newborns, *S. agalactiae*, *E. coli*, and *S. pneumoniae* are the predominant pathogens, while *N. meningitidis*, *H. influenzae*, and *S. pneumoniae* are the most common in children between 2 months and 16 years old (3). Conjugate vaccines against *S. pneumoniae*, *H. influenzae*, and *N. meningitidis* have reduced the incidence of this disease, but with replacement by non-vaccine pneumococcal serotypes and the emergence of bacterial strains with reduced susceptibility to antimicrobial treatment, BM has become a health challenge worldwide (4). Therefore, understanding of epidemiological patterns, clinical characteristics, pathogen distribution, and antimicrobial susceptibility is important for the development of effective public health policies.

The World Health Organization (WHO) has coordinated the Global Invasive Bacterial Vaccine-Preventable Disease (IB-VPD) Surveillance Network (GISN) since 2009. The network was created to standardize monitoring of the burden and etiology of IB-VPD and to determine the impact of the introduction of conjugate pneumococcal vaccines (5). Colombia is part of this initiative with the participation of the HOMI, Fundación Hospital Pediátrico la Misericordia, a tertiaryIZlevel private university hospital in Bogotá, Colombia (6, 7). The aim of this study was to describe the etiology, epidemiology, and clinical presentations of children under 5 years of age with bacterial meningitis at HOMI between 2016 and 2023.

## MATERIALS AND METHODS

### Design, population and study period

A cross-sectional study was carried out from January 1, 2016, to December 31, 2023, based on an active institutional search of medical records from children under 5 years of age with suspected BM. Epidemiological surveillance of BM followed the protocol established by the Pan American Health Organization (PAHO) (6). Any hospitalized child under five years of age with a clinical diagnosis of meningitis was considered a suspected BM case. These patients underwent blood culture and lumbar puncture. Based on laboratory findings, meningitis cases were subsequently classified as probable, confirmed, or ruled out. Probable cases of BM were defined as suspected cases of meningitis with cerebrospinal fluid (CSF) examination showing any of the following characteristics in the cytochemical analysis: turbidity or increased leukocytes (> 100/mm3), elevated proteins (> 100 mg/dL), decreased glucose levels (<40 mg/dl), or Gram stain with the presence of microorganisms and negative culture. A confirmed case of BM was defined as a probable case with identification by culture of a microorganism in CSF or blood culture (Vitek TM 2 (BioMerieux, Marcy I’Etoile, Francia) or by molecular tests (FilmArrayTM, Biofire, or real-time PCR). A discarded case was defined as a probable case in which the values of the CSF cytochemical analysis were not consistent with those of BM.

### Data collection

The clinical records of patients who met the definition of a suspected case were reviewed daily, and the demographic variables, clinical data, vaccination history, laboratory findings, and final patient condition were recorded in an Excel database. This information was reported in the New Vaccine Surveillance Base (https://www.paho.org/es/documentos/vinuva-ja2011-12).

### Laboratory procedures Bacterial isolates

At the sentinel institution, microorganisms isolated from blood cultures and CSF were identified using VITEK2 TM (bioMerieux, Marcy I’Etoile, France). Following the established protocols in the reference and counter-reference systems, the isolates were forwarded to the Public Health Laboratory of the Bogotá for confirmation using the VITEK 2 TM system (bioMérieux, Marcy l’Etoile, France). Subsequently, the samples were transferred to the Microbiology Group at the National Institute of Health (INS) for additional analyses. The isolates identified as *S. pneumoniae, H. influenzae* and *N. meningitidis* were serotyped by the Quellung reaction or slide agglutination using commercial antisera (DIFCO, Becton Dickinson) (8). Antimicrobial susceptibility profiles were determined using the disk diffusion test (Kirby-Bauer) and broth microdilution for the antibiotics penicillin (PEN), ampicillin (AMP), ceftriaxone (CRO), trimethoprim–sulfamethoxazole (SXT), chloramphenicol (CHL), tetracycline (TET), erythromycin (ERY), rifampicin (RIF), vancomycin (VA), and ciprofloxacin (CIP), considering the identified microorganisms. The results were interpreted according to the criteria of the current Clinical and Laboratory Standards Institute (CLSI) (9).

Since 2018, CSF with negative cultures from suspected BM patients were processed by FilmArray™ (BioFire™) at the sentinel institution and sent to the Microbiology Group - National Reference Laboratory (LNR) of the National Institute of Health for molecular testing to identify and serotyping/serogrouping of *S. pneumoniae*, *H. influenzae*, or *N. meningitidis* by PCR adapted from the US Centers for Disease Control and Prevention (Atlanta, Georgia), which included real-time PCR for the identification of *lyt*A (*S. pneumoniae*), *hpd* (*H. influenzae*), and *sod*C or *ctr*A (*N. meningitidis*) genes and multiplex PCR for serotyping/serogrouping (8).

### Selection bias

The risk of selection bias was assessed because the study was conducted in a tertiaryIZcare hospital that admits patients with more severe conditions. There was also a potential risk of bias due to loss of isolate viability during processing or shipment; therefore, a protocol for sample transport and viability verification was implemented.

### Statistical analysis

Data were entered into an Excel database to determine the frequency of demographic variables, clinical characteristics, and patient outcomes. The incidence of BM per year among children under five years of age was also analyzed.

## RESULTS

From January 1, 2016, to December 31, 2023, there were 527 suspected cases of BM among children under 5 years of age, of whom 22.8% (n = 120) complied with the definition of a probable case and 13.1% (n = 69) of these were confirmed as BM (Figure 1). The annual average number of suspected cases was 66 and of probable cases was 15. The year with the highest number of suspected cases (n=114), probable cases (n=33) and confirmed cases (n=17) was 2019. The same year also saw the highest incidence of BM among children under 5 years old, reaching 1.87 per 1,000 hospitalized children (Table 1).

**Figure 1.**
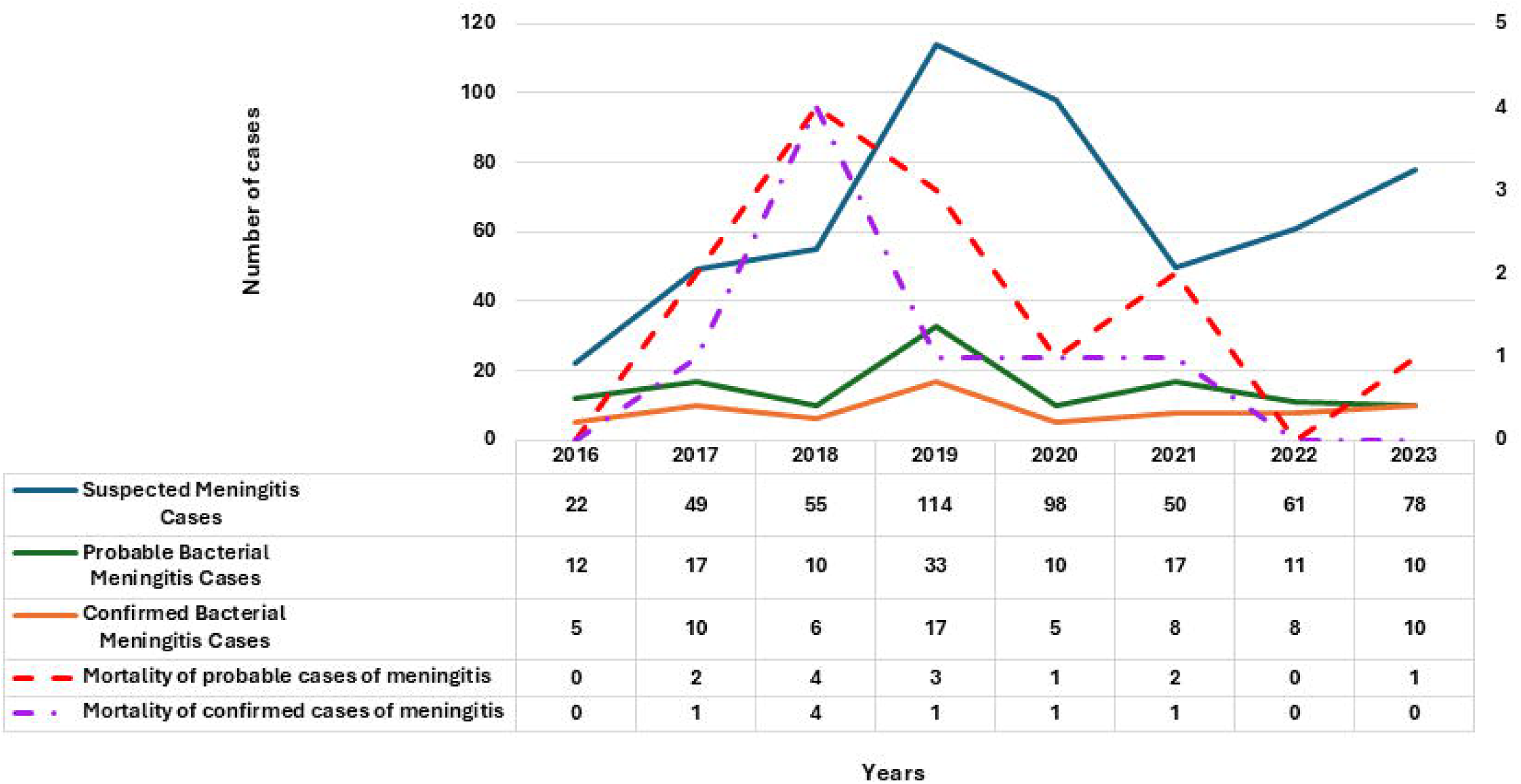
Number of BM cases in children under 5 years of age during sentinel surveillance at HOMI from 2016 to 2023.

**Table 1.** Demographic, clinical, and laboratory characteristics of confirmed bacterial meningitis cases and nonIZbacterial meningitis cases in children under 5 years of age, 2016–2023.

| Features | Classification of clinic cases |  | p value |
| --- | --- | --- | --- |
|  | Confirmed meningitis | Probable meningitis |  |
| Sex |  |  |  |
| Male | 41(59.4%) | 19(56.6%) | 0.42 |
| Female | 28(40.6%) | 22(43.4%) |  |
| Age Group (months) |  |  |  |
| < 12 | 48(69.6%) | 30(58.5%) | 0.11 |
| 12 - 23 | 6(8.7%) | 10(18.9%) | 0.16 |
| 24 - 35 | 4(5.8%) | 7(13.2%) | 0.18 |
| 36 - 47 | 7(10.1%) | 0(0,0%) | 0.21 |
| 48 - 59 | 4(5.8%) | 4(7.5%) | 1.00 |
| Cases | 69(57.5%) | 51(42.5%) | ---- |
| Median hospital stay (days) | 24.5 | 10.0 | ---- |
| Outcomes of patients |  |  |  |
| Death | 8(11.6%) | 5(9.8%) | 0.37 |
| Living with sequelae | 18(26.1%) | 5(9.8%) | 0.03 |
| Living without sequelae | 43(62.3%) | 41(80.4%) | 0.01 |
| Annual incidence x 1000 hospitalized children <5 years |  |  |  |
| 2016 | 0.56 | 0,79 | ---- |
| 2017 | 0.81 | 0.56 | ---- |
| 2018 | 0.58 | 0.39 | ---- |
| 2019 | 1.87 | 1.76 | ---- |
| 2020 | 0.94 | 0.94 | ---- |
| 2021 | 0.67 | 0.75 | ---- |
| 2022 | 0.57 | 0.21 | ---- |
| 2023 | 0.78 | 0.16 | ---- |
| Clinical symptoms |  |  |  |
| Severe malnutrition | 0(0.0%) | 1(2.0%) | 0.46 |
| Petechial/purple rash | 2(2.9%) | 1(2.0%) | 1.00 |
| Bulging of the fontanelle | 13(18.8%) | 4(7.8%) | 0.16 |
| Stiff neck | 15(21.7%) | 8(15.7%) | 0.60 |
| Difficulty drinking/breastfeeding | 15(21.7%) | 6(11.8%) | 0.13 |
| Prostration/lethargy | 19(27.5%) | 10(19.6%) | 0.19 |
| Seizures | 25(36.2%) | 20(39.2%) | 0.83 |
| Altered Consciousness | 27(39.1%) | 16(31.4%) | 0.23 |
| Fever ($\geq 38.5^{\circ}\text{C}$) | 60(98.3%) | 44(86.3%) | 0.79 |
| *Cerebrospinal fluid analysis* |  |  |  |
| White blood cell count > 100 cells/$\mu\text{L}$ | 38(55.1%) | 13(25.5%) | **<0.001** |
| Protein >100 mg/dL | 57(82.6%) | 33(64.7%) | 0.02 |
| Glucose <40 mg/dL | 41(59.4%) | 24(47.1%) | 0.21 |
| CSF appearance |  |  |  |
| Transparent | 11(15.9%) | 12(23.5%) | 0.64 |
| Cloudy | 49(71.1%) | 34(66.7%) | 1.00 |
| Bloody | 1(1.4%) | 2(3.9%) | 0.59 |
| Xanthochromic | 4(5.8%) | 1(2.0%) | 0.36 |
| Other | 4(5.8%) | 3(5.9%) | 1.00 |
| *Number of doses of PCV10 received* |  |  |  |
| 1 | 9(13.0%) | 5(9.8%) | 0.36 |
| 2 | 11(15.9%) | 13(25.5%) | 0.8 |
| 3 | 16(23.2%) | 17(33.3%) | 0.62 |
| *Number of doses of Hib vaccine received* |  |  |  |
| 1 | 8(11.6%) | 6(18.2%) | 0.55 |
| 2 | 4(5.8%) | 2(6.1%) | 0.35 |
| 3 | 25(36.2%) | 26(51.0%) | 0.19 |

Confirmed and probable cases of BM were more frequent among male patients (59.4% and 56.6%, respectively) and in children under 12 months of age (69.6% and 58.5%, respectively) (Table 1). The primary clinical symptoms observed in the two groups during the assessment were fever, altered consciousness, and seizures; however, these differences were not statistically significant (p > 0.05) (Table 1). The CSF laboratory test results indicated that leukocyte, protein, and glucose levels were slightly elevated in the confirmed cases group compared to the probable cases group, with statistical significance observed in the leukocyte counts above 100 cells/μL (p < 0.001) in BM patients. Among the patients with suspected BM, 9.8% experienced post-infection complications or died. In contrast, among those definitively diagnosed with BM, 26.1% developed sequelae, while 11.6% did not survive. Hospital stay was longer for patients with confirmed BM than for those with probable BM (24.5 days vs. 10 days). For the PCV10 vaccine, 57.7% of patients with confirmed BM and 60.0% of patients with probable BM reported receiving the vaccine according to recommended schedule. For the Hib vaccine, 73.3% of confirmed cases and 76.6% of probable cases had been vaccinated as documented on their vaccination card (Table 1). None of the patients had received meningococcal vaccine doses.

Among the confirmed cases of BM, *S. pneumoniae* was identified in 26.1% of the cases, *E. coli* in 21.7%, *H. influenzae* in 17.4%, *N. meningitidis* in 10.1%, and other microorganisms in 24.7% (Table 2). For *S. pneumoniae,* 10 (55.5%) isolates were viable for serotyping, and all corresponded to serotypes not included PCV10. For *H. infuenzae*, 75.0% were serotyped, 77.7% were serotype b (Hib), in children who had received age-appropriate vaccine doses, and the remaining 22.3% were classified as nonIZtypeable (HiNT). For *N. meningitidis* 71.4% were serotyped, of which 60.0% corresponded to serogroup C and 40.0% to serogroup Y (Table 3). These three microorganisms accounted to 65.2% of the etiological agents of BM in the study population. The antimicrobial resistance data of the viable isolates are presented in Table 3, which shows that most isolates were susceptible to all the antimicrobial agents evaluated. Lethality varied across pathogens: *S. pneumoniae* showed a lethality of 15.4% (2/13), *H. influenzae* 0% (0/11), *N. meningitidis* 16.7% (1/6), *E. coli* 8.3% (1/12), and other bacterial species 23.5% (4/17).

**Table 2.**
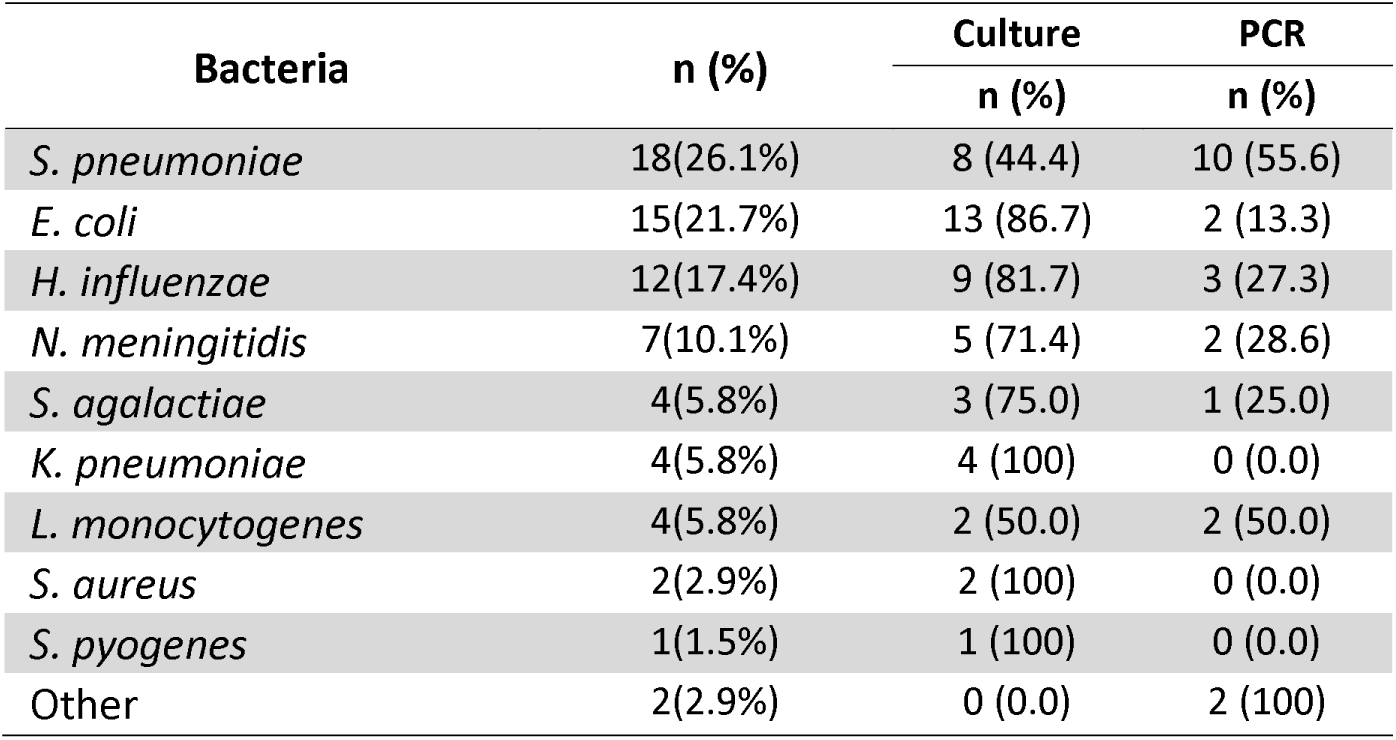
Microorganisms most frequently identified in cerebrospinal fluid and blood cultures from children under 5 years of age diagnosed with bacterial meningitis, 2016–2023.

| Bacteria | n (%) | Culture | PCR |
| --- | --- | --- | --- |
|  |  | n (%) | n (%) |
| *S. pneumoniae* | 18(26.1%) | 8 (44.4) | 10 (55.6) |
| *E. coli* | 15(21.7%) | 13 (86.7) | 2 (13.3) |
| *H. influenzae* | 12(17.4%) | 9 (81.7) | 3 (27.3) |
| *N. meningitidis* | 7(10.1%) | 5 (71.4) | 2 (28.6) |
| *S. agalactiae* | 4(5.8%) | 3 (75.0) | 1 (25.0) |
| *K. pneumoniae* | 4(5.8%) | 4 (100) | 0 (0.0) |
| *L. monocytogenes* | 4(5.8%) | 2 (50.0) | 2 (50.0) |
| *S. aureus* | 2(2.9%) | 2 (100) | 0 (0.0) |
| *S. pyogenes* | 1(1.5%) | 1 (100) | 0 (0.0) |
| Other | 2(2.9%) | 0 (0.0) | 2 (100) |

**Table 3.** Demographic characteristics, phenotypic features, and vaccine doses among confirmed cases of bacterial meningitis caused by *Streptococcus pneumoniae*, *Haemophilus influenzae*, and *Neisseria meningitidis* in children under 5 years of age, 2016–2023.

| Bacteria | Age group (months) | Serotype | Number of doses of Hib | Number of doses of PCV10 | Antimicrobial resistance<sup>a</sup> |
| --- | --- | --- | --- | --- | --- |
| *S. pneumoniae* | 24 - 35 | 42 | 3 | 3 | ERY |
|  | 12 - 23 | 15C | 2 | 2 | SXT |
|  | 48 - 59 | 15C | 3 | 3 | SXT |
|  | < 12 | 19A | 0 | 0 | No data<sup>b</sup> |
|  | 36 - 47 | 6A/6B/6C/6D | 3 | 3 | No data<sup>b</sup> |
|  | 48 - 59 | 6C/6D | 3 | 3 | No data |
|  | 36 - 37 | 23A | 3 | 3 | ERY |
|  | 36 - 47 | 23B | 3 | 3 | ERY |
|  | 36 - 47 | 23B | 3 | 3 | ERY |
|  | < 12 | 23B | 3 | 2 | Pansensible<sup>c</sup> |
| *H. influenzae* | < 12 | Hib | 2 | 2 | Pansensible |
|  | < 12 | Hib | 2 | 2 | No data |
|  | 48 - 59 | Hib | 3 | 0 | Pansensible |
|  | 48 - 59 | Hib | 3 | 0 | AMP, SXT |
|  | < 12 | Hib | 0 | 0 | AMP |
|  | < 12 | Hib | 0 | 0 | Pansensible |
|  | < 12 | Hib | 1 | 1 | Pansensible |
|  | < 12 | HiNT | 2 | 2 | Pansensible |
|  | < 12 | HiNT | 0 | 0 | Pansensible |
| *N. meningitidis* | < 12 | C | 1 | 1 | Pansensible |
|  | < 12 | C | 3 | 2 | No data |
|  | 12 - 23 | Y | 3 | 3 | Pansensible |
|  | 24 - 35 | C | 1 | 1 | PEN |
|  | < 12 | Y | 3 | 2 | PEN |

Note: No patient had vaccine data for *N. meningitidis*.
<sup>a</sup> Penicillin (PEN), ampicillin (AMP), ceftriaxone (CRO), trimethoprim-sulfamethoxazole (SXT), chloramphenicol (CHL), tetracycline (TET), erythromycin (ERY), rifampin (RIF), vancomycin (VA), and ciprofloxacin).
<sup>b</sup> Cases confirmed by PCR from cerebrospinal fluid and negative culture.
<sup>c</sup> Sensitive to all the antibiotics evaluated.

## DISCUSSION

BM continues to be a public health problem, with approximately 2.8 million new cases per year and a mortality rate of 21.0% worldwide (10). In 2021, the WHO published a route to combat bacterial meningitis by 2030, with an emphasis on vaccine-preventable pathogens, which is designed to improve diagnosis, strengthen meningitis surveillance systems, and increase access to vaccines to prevent infection (10). The present study describes the epidemiological, etiological, clinical, and laboratory characteristics of BM identified through sentinel surveillance in children under five years of age at a tertiaryIZcare hospital in Bogotá.

Surveillance of BM cases is essential for disease control and the vaccine development. Worldwide, in 2019, the incidence rate of meningitis in children under 5 years of age was 192.4 per 100,000 people (11). In Colombia, the annual incidence of BM in children is approximately 0.3-0.5 cases per 100,000 (12). During the study period, the highest hospital incidence observed in the sentinel institution was 1.87 per 1,000 hospitalized children in 2019. This highlights the importance of continuing surveillance of pediatric meningitis cases with laboratory-based confirmation of BM and identification of serotypes and serogroups, which is essential to detect the number of cases and identify the type of pathogen causing this deadly disease among children worldwide.

Among children under five years of age admitted to the hospital with suspected BM, 22.8% were classified as probable cases, while 13.1% were confirmed. The WHO, through the IB-VPD Network conducts surveillance of children under 5 years old admitted to sentinel hospitals for suspected meningitis. Data collected from 2014 to 2019 for this age group revealed that among the suspected meningitis cases, 8.6% were categorized as probable BM, whereas 2.6% were confirmed as BM (13). In Ghana, surveillance conducted at two sentinel sites revealed a frequency of 0.8% for confirmed cases (14). A multicenter hospital sentinel surveillance study involving 11 tertiary care facilities in India found that 4.5% of suspected cases were laboratory-confirmed as BM (15). The proportion of confirmed cases varies across regions due in large part to the complexity of the diagnosis of BM, implementation of vaccination programs for the prevention of infection, and differences in access to health systems. The proportion of confirmed cases in our study was higher than that in the other cited studies. This may be explained by the presence of a dedicated surveillance officer in the sentinel hospital, the implementation of a CO_2_ incubation system, access to molecular diagnostic methods either onIZsite or through a reference laboratory, and development of a strict protocol for the conservation and transportation of samples. In this study, the frequency of BM according to age and sex was similar to that observed in other prospective surveillance studies of hospital institutions in different geographic regions (13–15).

BM is associated with a poor prognosis, both in terms of mortality and long-term morbidity among survivors. In 2019, there were 236,000 deaths attributable to meningitis worldwide, 112,000 (87,400-145,000) were young children under 5 years of age (11), and the risk of disability due to sequelae in meningitis survivors after hospital discharge was estimated to be 20% (16). In this study, the mortality and sequelae rates were 11.6% and 26.1%, respectively. In a multicenter study conducted by 149 sentinel hospitals participating in surveillance sites in 58 countries and 6 countries, 11.0% of children with confirmed BM died and 5.1% were discharged from the hospital with sequelae (13), In a multicenter sentinel surveillance in India, the mortality rate was 8.7%, and the rate of development of post-infection sequelae was 5.0% (15). Regional variations in mortality and sequelae rates may be attributed to factors such as delayed effective antibiotic treatment, limited access to healthcare facilities, hospital care expenses, prevalent serotypes, the virulence of microorganisms, and antibiotic susceptibility profiles.

A systematic review revealed that *N. meningitidis*, *S. pneumoniae*, and *H. influenzae* remain the leading pathogens responsible for BM, with variations in pathogen prevalence based on age and geographic location (17). In our study cohort, 65.2% of BM cases were attributed to pathogens that could be prevented through vaccination. These findings are similar to data reported from other Latin American countries (18). Data from prospective research on bacterial meningitis showed regional differences in pathogen prevalence: *H. influenzae* was the main cause in Finland and Latin America, whereas *S. pneumoniae* was the most prevalent pathogen in Angola (19), and *N. meningitidis* was the most common pathogen among children under five years of age in Europe (17).

*S. pneumoniae* was the most frequent etiological agent (20.3%) of BM in children under five years of age, and the serotypes identified were different from those contained in PCV10. In Colombia, in 2009, PCV7 was introduced for free in the National Immunization Program of Colombia, which was replaced by PCV10 in January 2012 and PCV13 for populations born after May 1, 2022, in a 2 + 1 scheme at 2, 4, and 12 months of age. As a result of the introduction of PCV7/10 vaccines, postIZvaccination surveillance data in Colombia have shown a marked decline in PCV10-covered serotypes, accompanied by an increase in serotypes 19A, 3, and other non-PCV13 serotypes (20). Research conducted across 17 hospitals in Colombia examined meningitis cases in children caused by *S. pneumoniae*, showed that the proportion of serotypes covered by PCV10 dropped from 75% to 9.1% between 2016 and 2019, while the prevalence of vaccine serotypes 19A and 34 increased (21). This provides evidence that mass vaccination in Colombia has produced the expected impact, with a marked decline in cases caused by vaccineIZincluded serotypes and a concomitant increase in nonIZvaccine serotypes. Our study revealed that serotypes 23B, 15C, and 23A were the most identified serotypes. These serotypes have also emerged as the predominant strains carried by children and have been detected in invasive pneumococcal infections (22–24). This finding underscores the challenging scenario of serotype replacement following the implementation of pneumococcal conjugate vaccines. It also emphasizes the necessity for PCVs with broader coverage, at least until a universal vaccine that is not serotype-dependent becomes available.

*H. influenzae* accounted for 18.6% of the recovered isolates, with serotype b being the most prevalent. This serotype has been preventable through immunization in Colombia since the nationwide vaccination program began in 1998 (25). Hib cases were observed in children who had received full or partial vaccination (two doses) as well as in those who did not receive any dose. Research conducted in four Latin American countries indicated that high vaccination rates led to a reduced incidence of Hib meningitis (26). However, cases of vaccination failure have been documented in countries such as France (27), Brazil (28), and Argentina (29). The possible reappearance of Hib cases in countries with high vaccination coverage may be due to a decrease in collective protection due to suboptimal vaccination coverage rates, reduced antibody titers in children, vaccination schedules lacking booster doses, the appearance of more virulent strains and the use of different types of vaccines (27, 29, 30), which should be taken into account when formulating future strategies for the prevention, control and elimination of Hib. Based on these observations, in 2023, Colombia implemented a Hib booster (pentavalent vaccine) at 18 months of age. This strategy has proven effective in controlling the re-emergence of *H. influenza* serotype b infections in countries where the burden of infection affects older children (31).

Ten percent of the isolates were *N. meningitidis*. Universal meningococcal vaccination has not been implemented in Colombia as part of the national immunization programs; however, quadrivalent conjugate vaccines for serogroups A, C, Y, and W and a protein vaccine against meningococcus B (4CMenB) are available, and these vaccines are utilized in high-risk patients, sporadic outbreaks, and can be purchased at a cost to the user. A study to evaluate strategies against meningococcal disease found that in Colombia, providing universal immunization with two doses of Men-ACWY to infants older than six months is a cost-effective method based on the World Health Organization’s threshold of three times the gross domestic product (GDP). This strategy yielded an incremental cost-effectiveness ratio (ICER) of 16,191 per life-year saved (LYS) (32); therefore, vaccination to protect against these serogroups could be beneficial (33). In this study, *Escherichia coli* as a causative agent of BM was observed, with most cases occurring outside the neonatal period. These data coincide with publications that place it among the top five causes of bacterial meningitis in children under five years of age (16).

The strengths of this study include the coordinated participation of multiple institutions (HOMI, Fundación Hospital Pediátrico la Misericordia, Secretary of Health of Bogotá, National Institute of Health (INS), Ministry of Health and Pan American Health Organization), the prospective collection of information maintained during the COVID-19 pandemic , and the characteristics of the sentinel hospital, a level IV pediatric hospital, with the infrastructure and epidemiological support to ensure the quality of the data. However, this study has several limitations: (i) the study was conducted in a level IV hospital in Bogotá, and the data cannot be extrapolated to other hospital institutions or to the regional level. (ii) not all bacterial isolates from confirmed BM cases could be recovered; therefore, phenotypic characterization was not possible.

This study characterizes the demographic and clinical profile of BM in children under five years of age, confirming *S. pneumoniae*, *H. influenzae*, and *N. meningitidis* as the principal etiologic agents responsible for the disease. The findings also underscore the necessity of enhancing and maintaining sentinel epidemiological surveillance as it serves as a strategy that provides information on the behavior of preventable diseases through vaccination to take actions that contribute to the public health of the population.

## FUNDING SOURCE

Pan American Health Organization/World Health Organization

National Institute of Health

District Health Secretary of Bogotá

HOMI, Fundación Hospital Pediátrico la Misericordia.

## ETHICAL APPROVAL STATEMENT

This was a risk-free study since the patients were not subjected to any intervention, procedure or test other than those indicated for the diseases. It was approved by the Ethics and Research Committee of HOMI, Fundación Hospital Pediátrico la Misericordia by CEI 2015, 2015 act.

## AUTHORS’ CONTRIBUTIONS

CD, GG-M, JM: Conceptualization and design of the study, data collection, validation of information, statistical analysis, project administration, writing, review, editing and approval of the document. AB, OS, LYM, MPPR, ES, YMER: Data collection, Laboratory methods, supervision and validation of information, statistical analysis, formal analysis, writing, review, editing and approval of the document. DJ, JO, JCRS, MCD, KJ: Conceptualization, supervision and validation of information, statistical analysis, formal analysis, acquisition of resources, writing, review, editing and approval of the document.

## AUTHOR APPROVAL

All authors have seen and approved the manuscript.

## Data Availability

All data produced in the present study are available upon reasonable request to the authors.

## REFERENCES

1. Hasbun R. Progress and Challenges in Bacterial Meningitis: A Review. JAMA. 2022 Dec 6;328(21):2147–2154. doi: 10.1001/jama.2022.20521. Erratum in: JAMA. 2023 Feb 14;329(6):515. doi: 10.1001/jama.2023.0570. PMID: 36472590

2. Wang C, Xu H, Liu G, Liu J, Yu H, Chen B, et. Al. A Multicenter Epidemiological and Pathogenic Characteristics Study of Community-Acquired Bacterial Meningitis Children in China: Results from the Chinese Pediatric Bacterial Meningitis Surveillance (CPBMS) 2019-2020. Infect Drug Resist. 2023 Oct 9;16:6587–6601. doi: 10.2147/IDR.S413147. PMID: 37849791; PMCID: PMC10577658.

3. van Ettekoven CN, Liechti FD, Brouwer MC, Bijlsma MW, van de Beek D. Global Case Fatality of Bacterial Meningitis During an 80-Year Period: A Systematic Review and Meta-Analysis. JAMA Netw Open. 2024 Aug 1;7(8):e2424802. doi: 10.1001/jamanetworkopen.2024.24802. PMID: 39093565; PMCID: PMC11297475

4. van de Beek D, Brouwer MC, Koedel U, Wall EC. Community-acquired bacterial meningitis. Lancet. 2021 Sep 25;398(10306):1171–1183. doi: 10.1016/S0140-6736(21)00883-7. Epub 2021 Jul 22. PMID: 34303412.

5. Nakamura T, Cohen AL, Schwartz S, Mwenda JM, Weldegebriel G, Biey JNM, et al. The Global Landscape of Pediatric Bacterial Meningitis Data Reported to the World Health Organization-Coordinated Invasive Bacterial Vaccine-Preventable Disease Surveillance Network, 2014-2019. J Infect Dis. 2021 Sep 1;224(12 Suppl 2):S161–S173. doi: 10.1093/infdis/jiab217. PMID: 34469555; PMCID: PMC8409679.

6. Camacho-Moreno G, Duarte C, García D, Calderón V, Maldonado LY, Castellar L, et al. Sentinel surveillance for bacterial pneumonia and meningitis in children under the age of 5 in a tertiary pediatric hospital in Colombia - 2016. Biomedica. 2021 Oct 15;41(Sp. 2):62–75. doi: 10.7705/biomedica.5658. PMID: 34669279; PMCID: PMC8614369.

7. Camacho-Moreno G, Duarte C, Perdomo MDP, Maldonado LY, Palacios J, Rojas JC, et al. Sentinel surveillance in bacterial pneumonia in children under 5 years old in a fourth-level pediatric hospital in Colombia 2016-2022. IJID Regions 2024 (13): 100449. 10.1016/j.ijregi.2024.100449.

8. World Health Organization and Centers for Disease Control and Prevention. Laboratory methods for the diagnosis of meningitis caused by Neisseria meningitidis, Streptococcus pneumoniae, and Haemophilus influenzae: WHO manual. 2nd ed. https://apps.who.int/iris/handle/10665/70765. Accessed 15 January 2025

9. Performance Standards for Antimicrobial Susceptibility Testing: Twenty-sixth Informational Supplement Clinical Laboratory Standards Institute Performance Standards for Antimicrobial Susceptibility Testing: 33rd Edition. 2023.

10. Defeating meningitis by 2030: baseline situation analysis (Defeating meningitis by 2030: (who.int)), accessed January 16, 2025.

11. GBD 2019 Meningitis Antimicrobial Resistance Collaborators. Global, regional, and national burden of meningitis and it’s a etiologies, 1990-2019: a systematic analysis for the Global Burden of Disease Study 2019. Lancet Neurol. 2023 Aug;22(8):685–711. doi: 10.1016/S1474-4422(23)00195-3. PMID: 37479374; PMCID: PMC10356620

12. Pitre-Daza, A. D., Guerrero-Gómez, M. G., Gelvez-Ávila, R. E. (2023). Meningitis Bacteriana y sus Complicaciones a Corto Plazo en Niños de 0 a 5 Años en el Hospital Universitario Erasmo Meoz en el Lapso 2019-2021. Available at: https://repositorio.udes.edu.co/entities/publication/a5bf6fef-8909-4670-bd24-1baf12260cb7

13. Nakamura T, Cohen AL, Schwartz S, Mwenda JM, Weldegebriel G, Biey JNM, et al. The Global Landscape of Pediatric Bacterial Meningitis Data Reported to the World Health Organization-Coordinated Invasive Bacterial Vaccine-Preventable Disease Surveillance Network, 2014-2019. J Infect Dis. 2021 Sep 1;224(12 Suppl 2):S161–S173. doi: 10.1093/infdis/jiab217. PMID: 34469555; PMCID: PMC8409679

14. Renner LA, Usuf E, Mohammed NI, Ansong D, Dankwah T, Kusah JT, et al. Hospital-based Surveillance for Pediatric Bacterial Meningitis in the Era of the 13-Valent Pneumococcal Conjugate Vaccine in Ghana. Clin Infect Dis. 2019 Sep 5;69(Suppl 2):S89–S96. doi: 10.1093/cid/ciz464. PMID: 31505622; PMCID: PMC6736167

15. Jayaraman Y, Veeraraghavan B, Girish Kumar CP, Sukumar B, Rajkumar P, Kangusamy B, et al. Hospital-based sentinel surveillance for bacterial meningitis in under-five children prior to the introduction of the PCV13 in India. Vaccine. 2021 Jun 23;39(28):3737–3744. doi: 10.1016/j.vaccine.2021.05.041. Epub 2021 May 29. PMID: 34074545

16. Edmond K, Clark A, Korczak VS, Sanderson C, Griffiths UK, Rudan I. Global and regional risk of disabling sequelae from bacterial meningitis: a systematic review and meta-analysis. Lancet Infect Dis. 2010 May;10(5):317–28. doi: 10.1016/S1473-3099(10)70048-7. PMID: 20417414

17. Oordt-Speets AM, Bolijn R, van Hoorn RC, Bhavsar A, Kyaw MH. Global etiology of bacterial meningitis: A systematic review and meta-analysis. PLoS One. 2018 Jun 11;13(6):e0198772. doi: 10.1371/journal.pone.0198772. PMID: 29889859; PMCID: PMC5995389.

18. Informe regional de SIREVA II, 2017. Washington, D.C.: Organización Panamericana de la Salud; 2020. Licencia: CC BY-NC-SA 3.0 IGO. Available at: https://www.paho.org/sites/default/files/2021-02/sireva2017.jpg

19. Peltola, H., Roine, I., Kallio, M. et al. Outcome of childhood bacterial meningitis on three continents. Sci Rep 11, 21593 (2021). 10.1038/s41598-021-01085-w

20. National Institute of Health. Laboratory surveillance of Streptococcus pneumoniae in Colombia, 2016-202. Available at https://www.ins.gov.co/BibliotecaDigital/vigilancia-por-laboratorio-de-streptococcus-pneumoniae-en-colombia-2016-2021.pdf

21. Farfán-Albarracín JD, Camacho-Moreno G, Leal AL, Patiño J, Coronell W, Gutiérrez IF, et al. Changes in the incidence of acute bacterial meningitis caused by *Streptococcus pneumoniae* and the implications of serotype replacement in children in Colombia after mass vaccination with PCV10. Front Pediatr. 2022 Sep 23;10:1006887. doi: 10.3389/fped.2022.1006887. PMID: 36210950; PMCID: PMC9545348

22. Candeias C, Almeida ST, Paulo AC, Simões AS, Ferreira B, Cruz AR, et. al. *Streptococcus pneumoniae* carriage, serotypes, genotypes, and antimicrobial resistance trends among children in Portugal, after introduction of PCV13 in National Immunization Program: A cross-sectional study. Vaccine. 2024 Sep 17;42(22):126219. doi: 10.1016/j.vaccine.2024.126219. Epub 2024 Aug 14. PMID: 39146858.

23. Ben-Shimol S, Givon-Lavi N, Kotler L, Adriaan van der Beek B, Greenberg D, Dagan R. Post–13-Valent Pneumococcal Conjugate Vaccine Dynamics in Young Children of Serotypes Included in Candidate Extended-Spectrum Conjugate Vaccines. Emerg Infect Dis. 2021;27(1):150–160. 10.3201/eid2701.201178.

24. Desmet S, Wouters I, Heirstraeten LV, Beutels P, Van Damme P, Malhotra-Kumar S, et. al. In-depth analysis of pneumococcal serotypes in Belgian children (2015-2018): Diversity, invasive disease potential, and antimicrobial susceptibility in carriage and disease. Vaccine. 2021 Jan 8;39(2):372–379. doi: 10.1016/j.vaccine.2020.11.044. Epub 2020 Dec 9. PMID: 33308889

25. Rodríguez MK, Agudelo CI, Duarte C. Aislamientos invasivos de *Haemophilus influenzae* en menores de 5 años: distribución de los serotipos y de la sensibilidad antimicrobiana, SIREVA II, Colombia 2002-2013. Infectio. 2015 19(2), 67–74. 10.1016/j.infect.2014.12.005

26. Slack MPE, Cripps AW, Grimwood K, Mackenzie GA, Ulanova M. Invasive *Haemophilus influenzae* Infections after 3 Decades of Hib Protein Conjugate Vaccine Use. Clin Microbiol Rev. 2021 Jun 16;34(3):e0002821. doi: 10.1128/CMR.00028-21. Epub 2021 Jun 2. PMID: 34076491; PMCID: PMC8262803

27. Hong E, Terrade A, Denizon M, Aouiti-Trabelsi M, Falguières M, Taha MK, Deghmane AE. *Haemophilus influenzae* type b (Hib) seroprevalence in France: impact of vaccination schedules. BMC Infect Dis. 2021 Jul 30;21(1):715. doi: 10.1186/s12879-021-06440-w. PMID: 34330228; PMCID: PMC8325224

28. Tuyama M, Corrêa-Antônio J., Schlackman J, Marsh JW, Rebelo MC, Cerqueira EO, et al. Invasive *Haemophilus influenzae* disease in the vaccine era in Rio de Janeiro, Brazil. Memórias do Instituto Oswaldo Cruz, (2017) 112, 196–202

29. Efron A, Nápoli D, Neyro S, Juárez MDV, Moscoloni M, Eluchans NS, et al. Laboratory surveillance of invasive Haemophilus influenzae disease in Argentina, 2011-2019. Rev Argent Microbiol. 2023 Apr-Jun;55(2):133-142. doi: 10.1016/j.ram.2022.08.002. Epub 2022 Oct 11. PMID: 36229277

30. Slack M, Esposito S, Haas H, Mihalyi A, Nissen M, Mukherjee P, Harrington L. *Haemophilus influenzae* type b disease in the era of conjugate vaccines: critical factors for successful eradication. Expert Rev Vaccines. 2020 Oct;19(10):903–917. doi: 10.1080/14760584.2020.1825948. Epub 2020 Oct 10. PMID: 32962476

31. Slack, M. P. E. Long term impact of conjugate vaccines on *Haemophilus influenzae* meningitis: narrative review. Microorganisms, 2021 9(5), 886.

32. La Rotta Cepeda, Jorge Enrique. Cost-effectiveness analysis of alternate strategies for children immunization against meningococcal disease with quadrivalent conjugate vaccines in Colombia. 2020. Available in: https://repositorio.unbosque.edu.co/items/a4de2e98-b1d6-4a35-ab37-6902eb6331db. )

33. Coronell-Rodriguez W, Caceres DC, Cintra O, Guzman-Holst A. Epidemiology of Invasive Meningococcal Disease in Colombia: A Retrospective Surveillance Database Analysis. Infect Dis Ther. 2023 Dec;12(12):2709–2724. doi: 10.1007/s40121-023-00886-y. Epub 2023 Nov 15. PMID: 37966702; PMCID: PMC10746648.

